# Educational Browser-Native SIR Simulation: Analytical Benchmarks Showing Numerical Accuracy for Lightweight Epidemic Modeling

**DOI:** 10.64898/2026.04.15.26350961

**Authors:** JJ Ben-Joseph

## Abstract

Lightweight epidemic calculators are widely used for teaching and rapid scenario exploration, yet many omit the methodological detail needed for scientific reuse. We present a browser-native SIR calculator that exposes forward Euler and classical fourth-order Runge–Kutta (RK4) integration alongside epidemiologically interpretable outputs and a population-conservation diagnostic. The implementation is anchored to analytical properties of the deterministic SIR system, including the epidemic threshold, the peak condition, and the final-size relation. Benchmark experiments show that RK4 is essentially step-size invariant over practical discretizations, whereas Euler at a coarse one-day step overestimates peak prevalence by 3.97% and final size by 0.66% relative to a fine-step RK4 reference. These results demonstrate that browser-based tools can support publication-quality computational narratives when solver choice, diagnostics, and assumptions are treated as first-class outputs.

## 1 Introduction

Deterministic compartment models remain central to mechanistic reasoning in infectious-disease epidemiology because they make assumptions explicit and link interventions to interpretable dynamical quantities [1–4]. Among these models, the classical SIR system is especially valuable pedagogically because it is mathematically tractable, qualitatively rich, and simple enough to run in a browser without external software dependencies.

That accessibility creates a recurrent problem. Web calculators often present a curve and a few summary numbers while leaving unreported the numerical solver, time step, stability diagnostics, and assumptions required to evaluate whether the output is reproducible or even numerically trustworthy. In outbreak analysis, such omissions can be a major problem. Seemingly minor implementation decisions can distort peak timing, bias final-size summaries, or obscure the distinction between a mechanistic scenario and a calibrated forecast [5–7].

We details the computational mechanics behind the browser-native SIR calculator deployed on AgentCalc. Specifically, we (i) formalize the dynamical quantities the calculator should report and relate them to analytical properties of the continuous-time SIR system, (ii) document the actual browser implementation, including solver options, non-negativity handling, and conservation diagnostics, and (iii) provide concrete validation results and a concise reporting standard that allows calculator outputs to be used in other materials with minimal ambiguity.

## 2 Model formulation

### 2.1 State variables and governing equations

We consider the closed-population deterministic SIR model

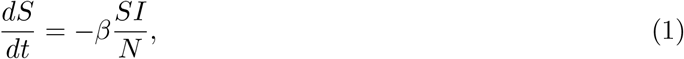

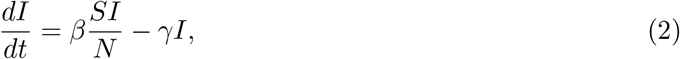

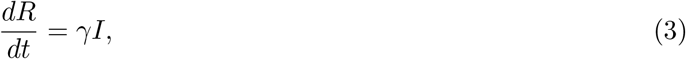

where *S*(*t*), *I*(*t*), and *R*(*t*) denote susceptible, infectious, and removed individuals, respectively, and *N* = *S*(*t*) + *I*(*t*) + *R*(*t*) is constant. The parameter *β* is the transmission rate and *γ* is the removal rate, so the mean infectious period is 1*/γ*.

To avoid the common notation clash between the initial removed compartment and the basic reproduction number, we write the initial removed count as *R*_init_ = *R*(0) and the basic reproduction number as

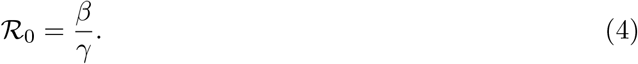

### 2.2 Derived epidemiological quantities

The calculator reports three summary families that are routinely interpretable across applications:

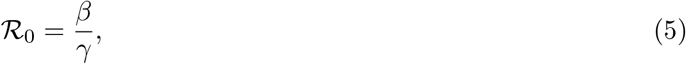

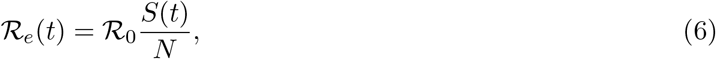

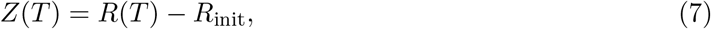

where ℛ_*e*_(*t*) is the instantaneous effective reproduction number under homogeneous mixing and *Z*(*T*) is the finite-horizon epidemic size. The browser interface displays ℛ_0_ together with the endpoint summaries ℛ_*e*_(0) and ℛ_*e*_(*T*); the full expression for ℛ_*e*_(*t*) is included here because it governs the growth-to-decline transition of the simulated trajectory. For a sufficiently long simulation horizon, *Z*(*T*) approximates the final epidemic size.

The infectious compartment evolves according to

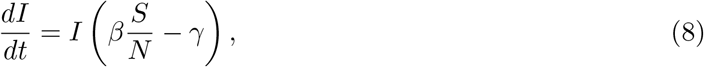

which makes the threshold structure explicit: *I*(*t*) initially grows when *S*(*t*) *> N/*ℛ_0_ and declines once *S*(*t*) *< N/*ℛ_0_. Therefore, in the continuous model, the prevalence peak occurs when

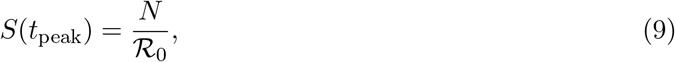

provided ℛ_0_ *>* 1 and an epidemic wave occurs [3, 8].

### 2.3 Final-size relation

The continuous SIR system also provides an analytical benchmark for terminal behavior. Dividing *dS* / *dt* by *dR* / *dt* yeilds

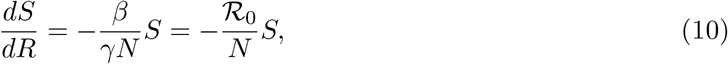

which integrates to

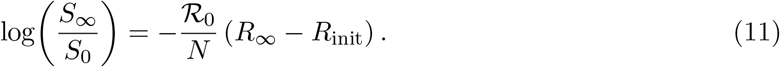

In the common case *R*_init_ = 0 and *S*_0_ ≈ *N*, the attack rate *z* = 1 − *S*_∞_*/N* satisfies the implicit final-size equation

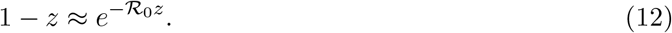

This relation is useful because it provides an implementation-independent check on long-horizon numerical results.

## 3 Browser implementation

### 3.1 Numerical integration schemes

The calculator exposes two explicit one-step solvers:

- **Forward Euler** 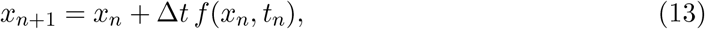

which is first-order accurate in the step size.
- **Classical RK4** 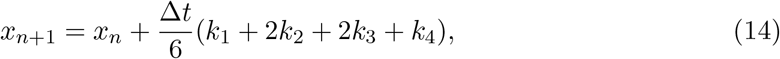

which is fourth-order accurate for smooth right-hand sides [9].

The browser form takes *N, I*_0_, *R*_init_, *β, γ*, Δ*t*, total step count *K*, and solver choice as direct inputs. The simulation horizon is therefore

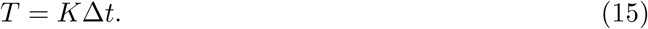

### 3.2 Practical safeguards and diagnostics

After each numerical step, the implementation truncates any negative compartment value to zero using a component-wise max(0, ·) rule before reporting results. This is a pragmatic user-interface safeguard: it prevents visually nonsensical negative populations from propagating into the displayed summaries. Scientifically, however, that safeguard should not be mistaken for numerical validity. A coarse step size can still be inappropriate even if the displayed state remains non-negative after truncation.

For this reason, the calculator reports a terminal population-conservation diagnostic

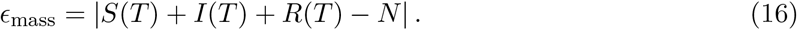

In a stable closed-population run, *ϵ*_mass_ should remain near floating-point roundoff. If clipping or step-size instability materially distorts the trajectory, the conservation diagnostic provides an immediately interpretable warning signal.

### 3.3 Reported outputs

The browser output panel returns:

1. simulation duration *T*,
2. terminal state (*S*(*T*), *I*(*T*), *R*(*T*)),
3. ℛ_0_ together with ℛ_*e*_(0) and ℛ_*e*_(*T*),
4. peak infectious prevalence max_*t*_ *I*(*t*),
5. time to peak,
6. finite-horizon epidemic size *Z*(*T*),
7. conservation error *ϵ*_mass_.

These are sufficient for a short methods section, a figure caption, or a supplement table if reported faithfully.

## 4 Validation design

We evaluated the implementation using tests that map directly onto the scientific claims of the calculator rather than only onto software behavior.

### 4.1 V1: Step-size convergence

Using the benchmark scenario *N* = 1000, *I*_0_ = 1, *R*_init_ = 0, *β* = 0.3 day^−1^, *γ* = 0.1 day^−1^, and *T* = 160 days, we compared Euler and RK4 trajectories across decreasing time steps. This scenario implies ℛ_0_ = 3 and is sufficiently nonlinear to expose solver differences.

### 4.2 V2: Peak-threshold consistency

For the same benchmark run, we checked whether the computed peak occurred near the analytical threshold *S* = *N/*ℛ_0_ = 333.33. Agreement between the numerical peak state and the threshold relation confirms that the browser implementation is respecting the underlying SIR geometry rather than merely producing a visually plausible curve.

### 4.3 V3: Conservation behavior

We summarized terminal *ϵ*_mass_ under the benchmark parameter set to distinguish ordinary floating-point error from implementation drift. Because the SIR equations conserve total population exactly in continuous time, any substantial deviation must be numerical.

## 5 Results

### 5.1 Benchmark accuracy

Table 1 reports representative outputs from the deployed browser algorithm. The fine-step RK4 run with Δ*t* = 0.05 days is used as a practical reference because further refinement changes displayed quantities only beyond the precision relevant to the interface.

**Table 1:**
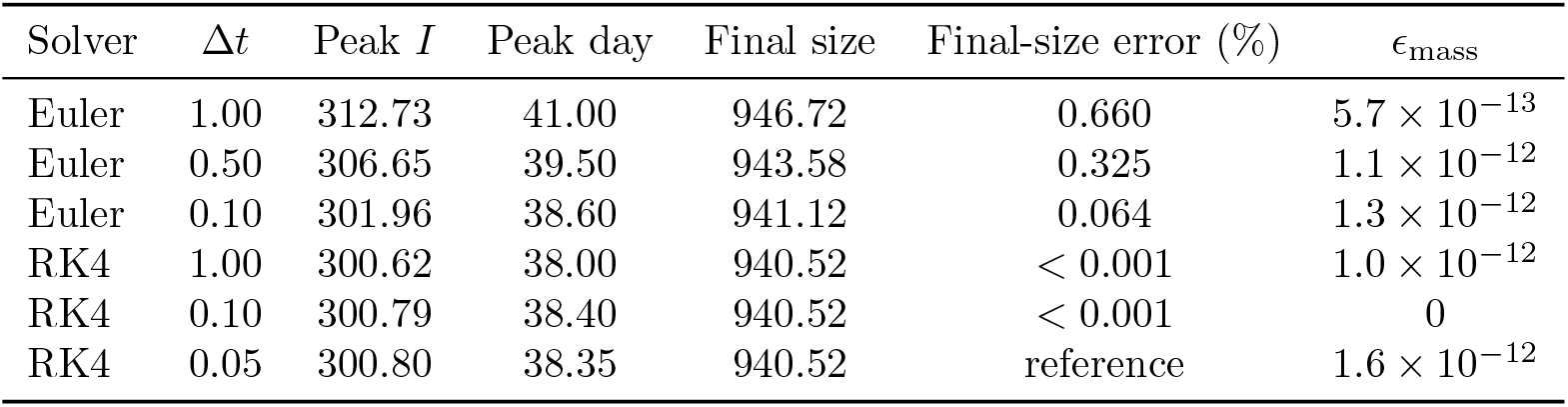
Representative solver and step-size benchmark for the browser implementation under *N* = 1000, *I*_0_ = 1, *R*_init_ = 0, *β* = 0.3, *γ* = 0.1, and *T* = 160 days. Relative final-size error is computed against the RK4 Δ*t* = 0.05 reference.

**Table 2:**
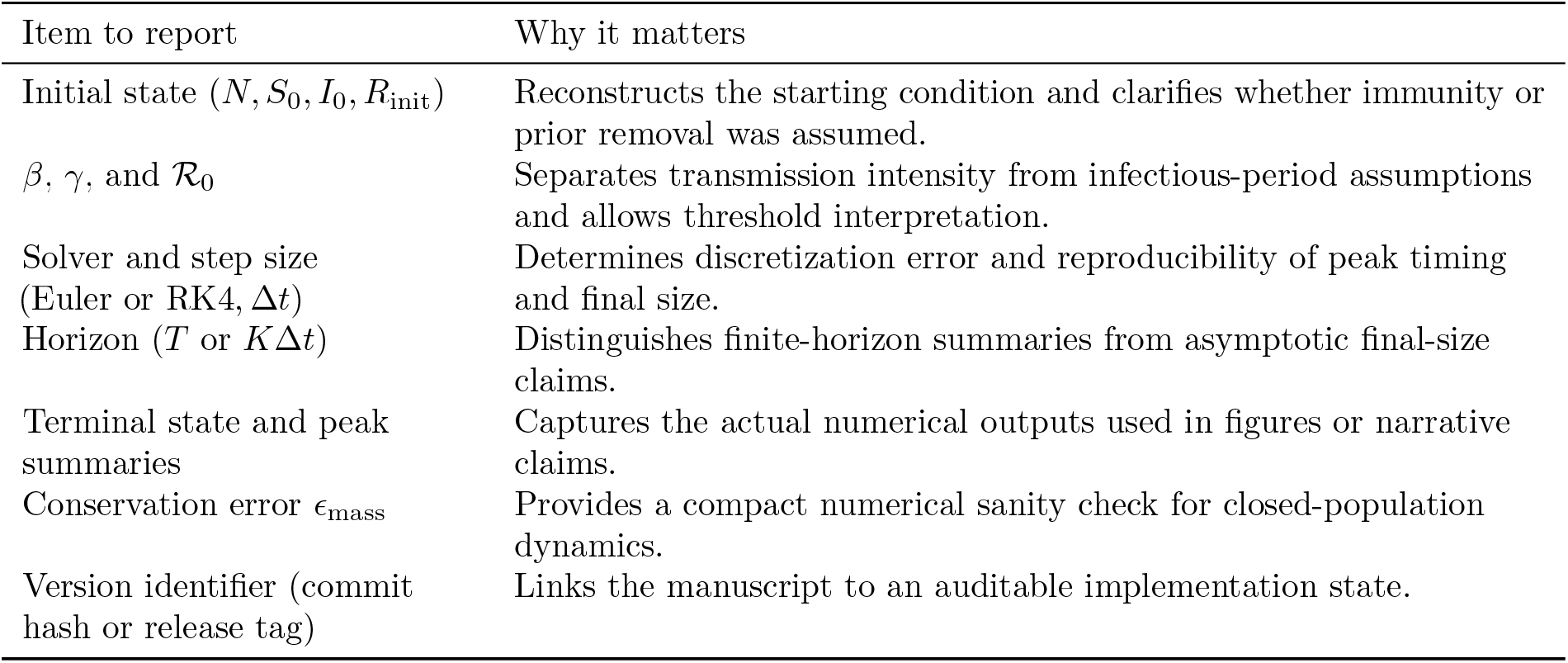
Minimal reporting standard for transferring browser-native SIR simulations into manuscripts or supplements.

Two features are immediately important. First, RK4 is nearly step-size invariant even at Δ*t* = 1 day for this benchmark, indicating that the higher-order method is robust enough for routine exploratory use when users prefer coarse inputs. Second, Euler converges monotonically toward the RK4 solution as Δ*t* decreases, but at a coarse one-day step it materially overstates both the epidemic peak and the cumulative number infected. That discrepancy is large enough to alter qualitative interpretation in educational or policy-adjacent discussions. Figure 1 makes the same point visually: the benchmark trajectory is smooth and internally consistent, yet the solver-step choice still changes the inferred burden near the peak.

**Figure 1:**
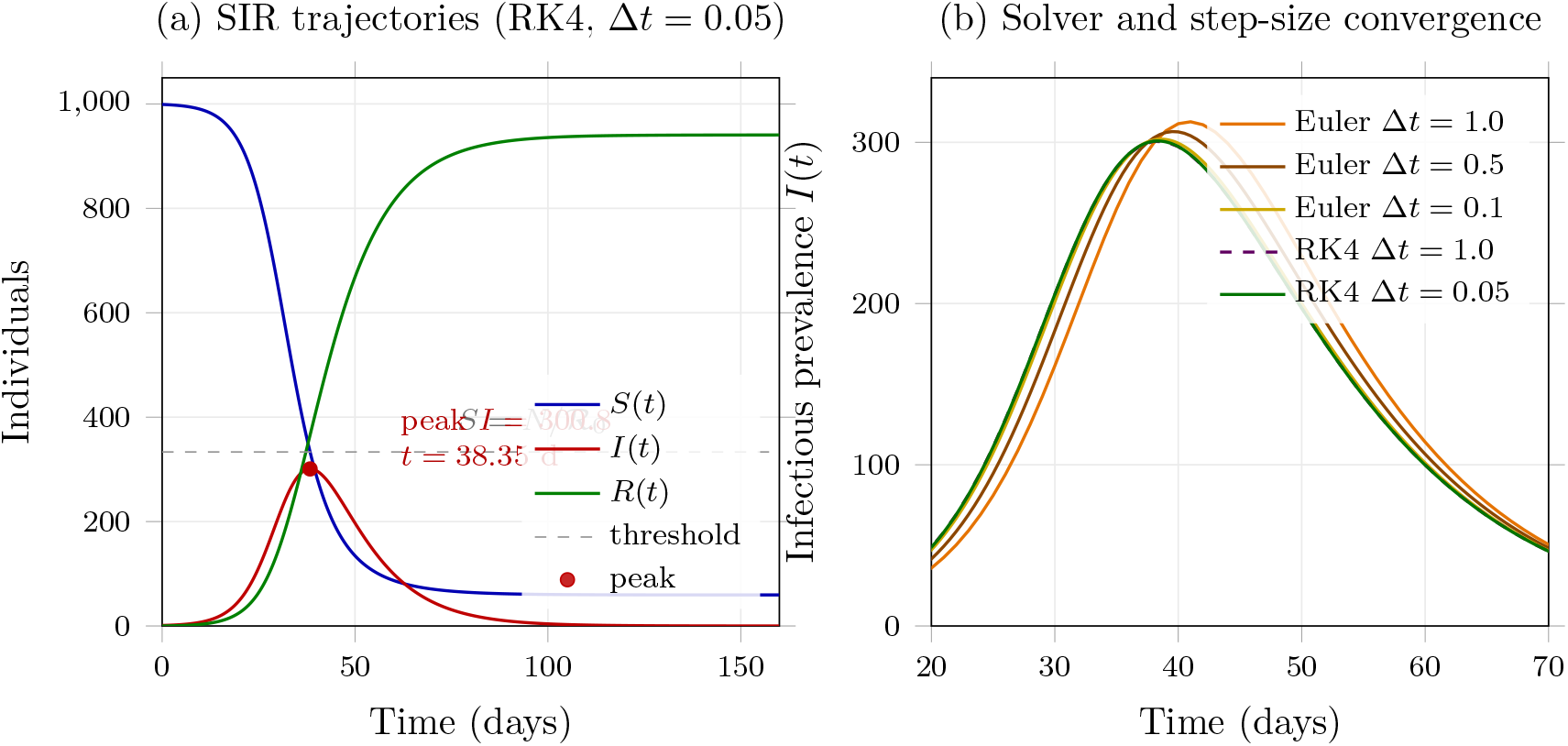
Benchmark behavior of the browser-native SIR implementation under *N* = 1000, *I*_0_ = 1, *R*_init_ = 0, *β* = 0.3, *γ* = 0.1, and *T* = 160 days. Left: reference RK4 trajectories at Δ*t* = 0.05 with the analytical peak threshold *S* = *N/*ℛ_0_ marked. Right: infectious-prevalence curves for representative Euler and RK4 step sizes, showing the coarser Euler discretization converging toward the RK4 reference as Δ*t* decreases.

### 5.2 Consistency with analytical benchmarks

For the RK4 reference run, the simulated prevalence peak occurred at day 38.35 with *S*(*t*_peak_) = 333.49, which is very close to the analytical threshold *N/*ℛ_0_ = 333.33. This agreement is scientifically more informative than a purely visual curve check because it confirms that the numerical implementation respects the defining peak condition of the SIR model.

The benchmark terminal susceptible count was *S*(*T*) ≈ 59.45, implying an attack rate near 94%. That magnitude is consistent with the final-size equation for ℛ_0_ = 3, reinforcing that the browser output lies on the expected SIR manifold rather than being an artifact of numerical drift.

### 5.3 Interpretation of the conservation diagnostic

Across the stable benchmark runs in Table 1, *ϵ*_mass_ remained at or below machine-precision scale. This is the desired regime. Importantly, however, the conservation diagnostic is necessary but not sufficient: an explicit solver can preserve the total population well while still shifting peak timing or final size. The benchmark therefore supports a practical rule for publication use: always report both the solver-step configuration and the conservation error.

## 6 Discussion

The main scientific value of a browser-native SIR tool is that it makes epidemic modeling easier and further it can make a mechanistic argument inspectable. When a calculator reveals the solver, the step size, the threshold quantities, and the diagnostic checks, it becomes possible to distinguish dynamical insight from merely presentational output.

Several practical lessons follow from the benchmark results. First, method choice matters even in apparently simple systems. Euler is perfectly serviceable for teaching numerical concepts and for small enough Δ*t*, but users should not assume that a one-day step is innocuous simply because the output curve looks smooth. Second, numerical clipping is a user-interface convenience, not a substitute for convergence analysis. If clipping is triggered under aggressive step sizes, the trajectory may look epidemiologically plausible while silently deviating from the intended model equations. Third, population conservation alone cannot certify inferential reliability. A simulation may conserve *N* almost perfectly while still shifting the epidemic peak by days.

These considerations matter especially when users move from exploration to inference. The present calculator is a forward simulator. It should not be used as a data-assimilation engine, an uncertainty-quantification framework, or a decision-support system. Fitting *β* and *γ* to noisy incidence data introduces non-identifiability, observation-process bias, and uncertainty propagation issues that require more formal statistical machinery [5, 6, 10]. Browser-native transparency is therefore most valuable at the stage of model communication, sensitivity analysis, and didactic exposition.

## 7 Limitations

The simulator intentionally preserves the minimal classical SIR assumptions: homogeneous mixing, no latent period, no demographic turnover, no age or network structure, no stochasticity, and time-invariant parameters. Those assumptions are often violated in operational settings [4, 11]. Consequently, the calculator should be used to reason about mechanisms and parameter sensitivity, not to make overconfident forecasts about real outbreaks.

The implementation also does not provide uncertainty intervals, structural-model comparisons, or calibration against surveillance data. An obvious next step is to pair the current reporting framework with an SEIR comparison mode or a parameter-sweep module so users can quantify how structural assumptions alter peak timing and final size.

## 8 Conclusion

A browser-native epidemic simulator can meet a publishable standard if it is explicit about model structure, numerical method, diagnostics, and inferential scope. In the present implementation, RK4 provides strong accuracy across practically useful step sizes, whereas Euler requires finer discretization to avoid meaningful bias in peak and cumulative summaries. More broadly, the work shows that lightweight web tools can support serious computational epidemiology narratives when reproducibility and discussion are designed into the output rather than retrofitted after the fact.

## Data Availability

https://github.com/tensorspace-ai/agentcalc_science

## Data and code availability

Versioned source files for the calculator, manuscript, and figure-generation script are available at https://github.com/tensorspace-ai/agentcalc_science. No human-participant data were collected or analyzed for this study.

